# Genomic characterization of a lytic bacteriophage targeting multidrug resistant *Klebsiella pneumoniae* from hospital sewage in Dhaka, Bangladesh

**DOI:** 10.1101/2023.08.13.23294043

**Authors:** Fahad Khan, Fariza Shams, Mohammad Sami Salman Bhuiyan, Aura Rahman, Syeda Naushin Tabassum, Md. Sakib Abrar Hossain, Abdus Sadique, Abdullah All Jaber, Jahidul Alam, Kamruzzaman Rumman, Probaha Biswas, Pronoy Debnath, Lovleen Tina Joshi, Sudhakar Bhandare, Maqsud Hossain

## Abstract

The emergence of multidrug-resistant (MDR) and extremely drug-resistant (XDR) *Klebsiella pneumoniae* presents a significant challenge to public health, particularly in hospital settings where it is a leading cause of nosocomial infections. Addressing the urgent need for alternative treatments to combat antibiotic-resistant bacteria, this study describes the isolation, characterization, and genomic analysis of a novel bacteriophage, designated as MFS, targeting MDR/XDR *K. pneumoniae* strains isolated from hospital sewage in Dhaka, Bangladesh. The phage was isolated utilizing a double-agar overlay and characterized using Oxford Nanopore Technologies sequencing. Phage MFS was identified as a member of the Siphoviridae family, under the unclassified Webervirus subfamily, with potent lytic activity against clinical MDR/XDR *K. pneumoniae* strains. Genomic analysis revealed a 48,780 bp genome with 94 protein-coding sequences, including essential genes for phage replication, structure, and host lysis, but notably lacking genes associated with antimicrobial resistance, virulence, and lysogeny. The presence of specific genes for endolysin and holin underscores the phage’s lytic capability. Additionally, the study elucidates the phage’s structural proteins and mechanisms underlying bacterial cell wall degradation, contributing valuable insights into phage-host interactions and applications of phage therapy. Our findings underscore the therapeutic potential of sewage-derived bacteriophages against MDR/XDR clinical *K. pneumoniae* and emphasize the need for further exploration of bacteriophage therapy as a viable alternative to traditional antibiotics in combating antibiotic-resistant bacterial infections.

## Introduction

*Klebsiella pneumoniae* (Kp) is a Gram negative, facultative anaerobic bacterium that belongs to the genus Klebsiella and the order *Enterobacterales*. *K. pneumoniae* is most commonly found in the human body’s upper respiratory tract and intestines [1]. It is one of the most common pathogens responsible for nosocomial infections, which include urinary tract infections, pneumonia, septicemia, and soft tissue infections [2], primarily in the elderly, infants, and immunocompromised patients. Initially, it was the extended-spectrum β-lactamases (ESBL) resistant *K. pneumoniae* strains that were the most common drug-resistant strains of *K. pneumoniae*. However, due to the rise in use of carbapenems, Carbapenem-resistant *K. pneumoniae* (CRKP) has been on the rise. As a result, carbapenem resistance in *K. pneumoniae* is frequently associated with antimicrobial resistance to other antibiotic classes, making treatment of infections challenging [3]. The Center for Disease Control and Prevention (CDC) and the European Center for Disease Prevention and Control have classified bacteria into MDR, extensively drug-resistant (XDR), and pan drug-resistant (PDR) categories [4]. Many *K. pneumoniae* strains are classified as XDR and are rapidly emerging as PDR by acquiring resistance to tigecycline and polymyxin antibiotics [5]. Antimicrobial resistance (AMR) is a major problem in Asia, and, recently, Bangladesh has shown a high prevalence of carbapenem-resistant organisms [6]. Among the carbapenemase genes, NDM-producing strains have been detected along with oxacillinase-48-like producing strains [3]. The emergence of hypervirulent and antibiotic-resistant strains emphasizes the critical need for effective anti-pathogen strategies to prevent its spread.

Bacteriophage therapy is a promising treatment option for antibiotic-resistant bacterial infections. Bacteriophages, also known as phages, are naturally occurring bacterial viruses that can specifically infect and lyse bacteria [7]. Phage therapy has gained renewed interest due to the growing problem of antimicrobial resistance and the lack of effective treatment options [10]. Studies on animal models have shown the efficacy of bacteriophages in treating infections caused by *K. pneumoniae*. For instance, in mouse models, bacteriophage therapy protected against respiratory and other infections caused by *K. pneumoniae*, such as liver abscesses and bacteremia [8, 9]. These promising findings highlight the potential of bacteriophage therapy as an effective alternative to antibiotics for the treatment of *K. pneumoniae* infections.

In this study we investigated the genome sequence of a *K. pneumoniae* bacteriophage that was isolated from a hospital sewage drain located at the Evercare Hospital in Dhaka, Bangladesh. The phage was named *Klebsiella* phage MFS (Accession Number: OP611406) and was propagated using K. *pneumoniae* strains that were previously collected from hospital samples from different regions in Dhaka, Bangladesh [10]. The isolation of phages from hospital sewage samples highlights the potential of these sources as rich and accessible reservoirs of bacteriophages that can be utilized for the development of novel therapeutics against medically significant pathogens such as *K. pneumoniae*.

## Methodology

### Bacterial strains and antibiotic susceptibility assay

Five clinical strains of *K. pneumoniae* (NGCE-327, NGCE-637, NGCE-639, NGCE-706, NGCE-707) were used in this study. All these strains were isolated from different clinical samples, including blood (n=2), urine (n=2), and sputum (n=1), originally collected from two tertiary care hospitals, Dhaka Central International Medical College & Hospital (DCIMCH) and Shaheed Suhrawardy Medical College and Hospital (SSMCH) (Supplementary File 1 S1). The collection dates for the isolates are as follows: NGCE-637 and NGCE-639 were collected on 04^th^ April 2017, NGCE-327 was collected on 17^th^ November 2018, and NGCE-706 and NGCE-707 were collected on 27^th^ February 2019. All clinical isolates were stored at −80°C in our lab.

The antibiotic susceptibility of the isolates was evaluated against amikacin, amoxicillin, cefepime, cefixime, ceftriaxone, cefuroxime, chloramphenicol, ciprofloxacin, colistin, gentamicin, levofloxacin, meropenem, nitrofurantoin, piperacillin/tazobactam, polymyxin B, trimethoprim, and tetracycline. Additionally, phenotypic screening for ESBL production was performed using the standard disk diffusion method. The results of the antibiotic susceptibility test were interpreted according to the CLSI (Clinical and Laboratory Standards Institute) guidelines [11]. Colistin susceptibility was determined using the standard broth microdilution method. Cation-adjusted Mueller Hinton Broth (HiMedia, India) was employed, and CLSI interpretive criteria (2020) were applied (S ≤ 2 mg/L, R > 2 mg/L).

### Bacteriophage isolation

In order to isolate lytic bacteriophage specific to *K. pneumoniae*, water sample was collected from a sewage outlet near tertiary care hospitals in Dhaka, Bangladesh. Furthermore, water samples were centrifuged (10,000 × g, 10 min) to remove solid particles. The supernatant was filtered through a 0.2 μm filter to exclude any bacteria present in the sample.

For the amplification of bacteriophage specific for *K. pneumoniae*, 250 ml of Luria Bertani (LB) broth and 10 ml of mid-log phase culture of *K. pneumoniae* strains (NGCE-327, NGCE-637, NGCE-639, NGCE-706, NGCE-707) and 1 ml phage filtrate were mixed and incubated for 24 hours at 37°C and 120 rpm in a shaker incubator. Following incubation, the flask contents were centrifuged for 10 minutes at 10000 pm. The supernatant was sterile filtered (0.2µm filter) and stored at 4° C. Single plaques were picked and purified three times with their corresponding hosts to obtain a pure phage using a double-layer agar method. The purified phages were stored at 4°C for further experiments [12].

The phage host range plays a crucial role in the effectiveness of phage therapy. Therefore, we wanted to assess the effect of the isolated phage on different MDR *Klebsiella* strains from our laboratory collection following the agar overlay assay as described elsewhere [13]. After overnight incubation at 37°C, the plates were observed for the presence of plaques on the bacterial lawns. The presence of plaque proves the lytic effect of phages on bacteria. The results were recorded as either lytic plaque, plaque with halo, turbid plaque or no plaque formation (68).

### Extraction of phage genomic DNA and library preparation for sequencing

Phage DNA was extracted using the Favorgen Viral DNA/RNA Kit (Taiwan). DNA concentrations were measured using a NanoDrop spectrophotometer, Model ND-1000 (Origin: Thermo Fisher Scientific, USA).

A Rapid PCR Barcoding Kit (SQK-RPB004) from Oxford Nanopore Technologies (Minion Mk1C) was used for library preparation. The library preparation protocol used was the simplest library preparation protocol that allowed sequencing from low DNA input and uses PCR to enable enough data to be generated. The process began with adding Fragmentation mix and barcoding with rapid attachment primer using PCR. After PCR, the products were cleaned using the

Agencourt AMPure XP, and rapid 1D sequencing adapters were attached to DNA ends, and the DNA was ready to be loaded on the primed flow cells. Finally, the libraries were sequenced using Minion Mk1C R9.4 flowcells, and real-time base calling was performed to generate fast files. The necessary reagents and flow cells were sourced from ONT (Littlemore, Oxford, United Kingdom).

### Genome Assembly

The raw FASTQ files were assembled using the Flye v2.9 assemblers for *de novo* assembly with default parameters to obtain high-quality contigs suitable for downstream analysis [14]. The contigs were then polished using Racon v1.4.20 [15] with the MinION reads as the consensus.

The assembled contigs were mapped back to the reference bacteriophage using Contiguator v2.7 [16], and the full-length contigs of the bacteriophage were identified. The contigs fasta files were used for downstream bioinformatics analysis, which included gene prediction, annotation, and functional analysis. The average nucleotide identity (ANI) of the phage was determined using OrthoANIb (OrthoANI using BLAST) [17]. The genome with an ANI >95% was designated as the same species. Furthermore, functional annotation was performed using the Rapid Annotation using the Subsystem Technology (RAST) server [18].

### Genomic Analysis

The genome of the phage was annotated using Prokka [19] and PHASTER [20], and a comprehensive diagram was generated using SnapGene® software (GSL Biotech; available at snapgene.com). The phage taxonomy was determined by adding the phage isolate genome to the VIP-tree database [21] and performing BLASTn and tBLASTn analysis against NCBI [22]. Codon usage frequencies in the phage genomes were computed using Codon Usage programme (https://www.bioinformatics.org/sms2/codon_usage.html) [23]. The presence of tRNAs was predicted using ARAGORN and tRNAscan-SE tools [24, 25]. The rho–independent transcription terminators were determined using ARNold (http://rssf.i2bc.paris-saclay.fr/toolbox/arnold/index.php). Possible anti-CRISPR genes were predicted using the AcrDB tool (https://bcb.unl.edu/AcrFinder) [26]. The presence of virulence determinants was screened using Virulence Factor Database (VFDB) [27] while antimicrobial resistance and temperate genetic signatures were determined using Comprehensive Antibiotic Research Database (CARD) [28]. Signal peptides were predicted by DeepTMHMM (v1.0.12) and SignalP-6.0 (v0.0.52) [29]. Phyre 2 (http://www.sbg.bio.ic.ac.uk/~phyre2) was used to predict the secondary structure of the proteins [30].

## Results

### Antibiotic Susceptibility profiles of *K. pneumoniae* Isolates

Based on the antibiotic susceptibility profiles, three isolates (NGCE-637, NGCE-639, NGCE-706) were multidrug resistant (MDR), and two strains (NGCE-707 and NGCE-327) classified as extensively drug-resistant (XDR). Across the spectrum, the antibiotic susceptibility testing results indicated universal resistance to amoxicillin, cefepime, cefixime, ceftriaxone, cefuroxime, nitrofurantoin, trimethoprim, and tetracycline. Except for one strain, NGCE-639, the other four strains were susceptible to colistin (Supplementary File 1 S1). These findings underscore the emergence of multidrug-resistant *Klebsiella pneumoniae* strains, exhibiting resistance across multiple antibiotic classes, including critical ones like carbapenems. The presence of ESBL enzymes in these isolates further exacerbates their resistance profile. Hence, bacteriophages can offer an alternative treatment for *K. pneumoniae* infections.

### Isolation of bacteriophage

Using *K. pneumoniae* NGCE 706 as an indicator bacterium, a lytic phage, *Klebsiella phage mfs*, was isolated from hospital sewage water in Dhaka, Bangladesh. This is the first report of a phage from this source. The lytic spectrum of the phage MFS extended to four other *K. pneumoniae* strains. The phage MFS produced plaques on the lawn of *K. pneumoniae* strains, three of which NGCE-637, NGCE-639, NGCE-706 was MDR, and two strains (NGCE-707 and NGCE-327) were XDR. The morphology of plaques was either completely clear or halo with a central transparent zone (Fig 1).

**Fig. 1.**
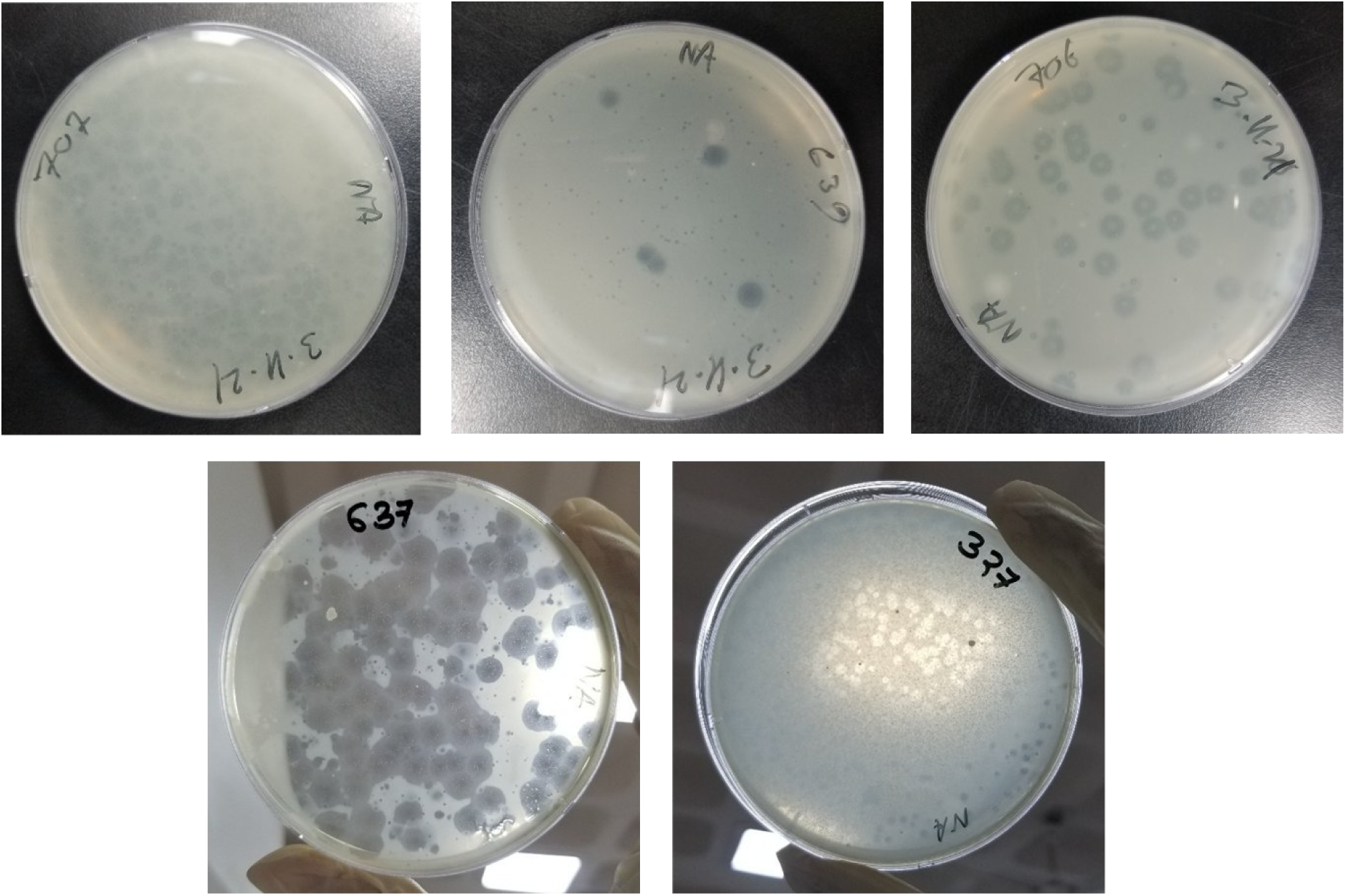
Plaque morphologies of the phage MFS on bacterial lawns of different *K. pneumoniae*. Plaques were allowed to develop in double layer agar plates overnight at 37 ◦C

Moreover, bacteriophages have shown exceptional efficacy against both MDR and XDR strains of *K. pneumoniae*, even when obtained from diverse sources like urine, blood and sputum. Previous studies have shown a high mortality rate in *K. pneumoniae* infections. [31]. Hence, phages can effectively battle both MDR and XDR Klebsiella isolates by entering biofilms, breaking bacterial cell walls, and interfering with bacterial replication processes [32].

### Genome Sequencing and Sequence Analysis

The complete genome sequencing of the *Klebsiella* phage MFS was carried out using the Oxford Nanopore MinION platform. The assembled read was found to have a size of 48,780 base pairs (bp) with a GC content of 52.02% (Table 1). The genome was linear and intact, and the whole genome sequence of *Klebsiella* phage MFS (Accession Number: OP611406) was obtained.

**Table 1.**
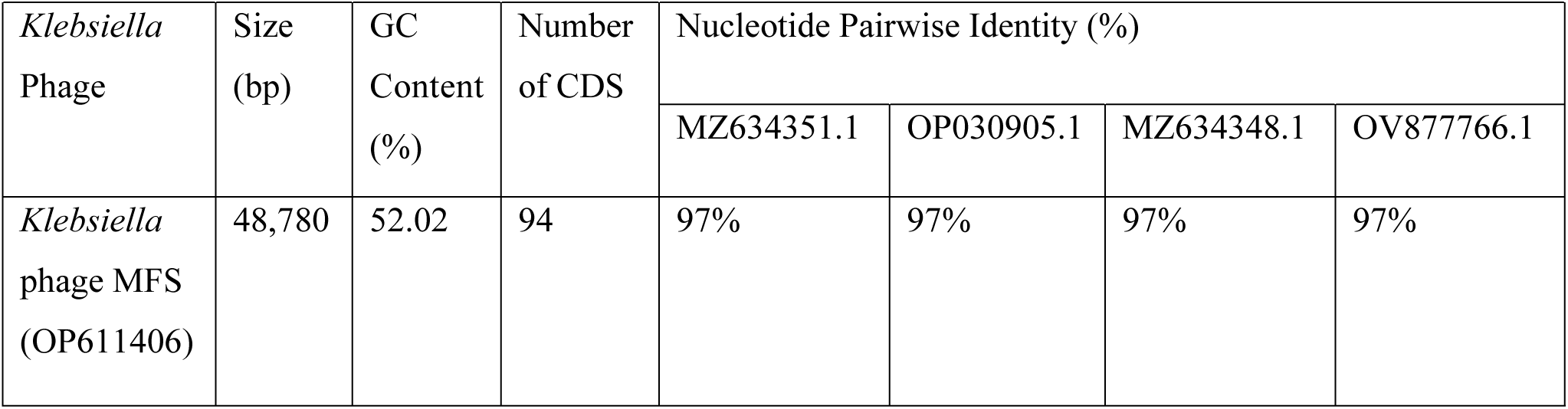
*De novo* assembly statistics of the novel *Klebsiella* phage MFS genome.

| *Klebsiella*<br>Phage | Size<br>(bp) | GC<br>Content<br>(%) | Number<br>of CDS | Nucleotide Pairwise Identity (%) |  |  |  |
| --- | --- | --- | --- | --- | --- | --- | --- |
|  |  |  |  | MZ634351.1 | OP030905.1 | MZ634348.1 | OV877766.1 |
| *Klebsiella*<br>phage MFS<br>(OP611406) | 48,780 | 52.02 | 94 | 97% | 97% | 97% | 97% |

Sequence analysis verified that the Phage MFS is assigned to the class *Caudoviricetes*, phylum *Uroviricota*, and unclassified *Webervirus* genus within the *Drexlerviridae* family. All tailed bacterial and archaeal viruses with icosahedral capsids and double-stranded DNA genomes belong to the class *Caudoviricetes*, which also includes members of the former morphology-based families *Myoviridae*, *Podoviridae*, and *Siphoviridae* [33].

A Megablast search of phage MFS genome indicated it has high sequence similarity to *Klebsiella* phage PWKp20, complete genome (96.78%), *Klebsiella* phage PWKp17(96.52%), *Klebsiella* phage PWKp19 (96.58%) (Table 1). According to Megablast search, MFS phage was placed in an unclassified webevirus under Drexlerviridae family.

The use of BLAST analysis further identified that these *Klebsiella* phages were part of the genus Webervirus (as shown in Fig 2). Multiple sequence alignment revealed that *Klebsiella* phage MFS was collinear and highly conserved compared to other weberviruses, specifically with phages *Klebsiella* phage PWKp20 (Accession Number: MZ634351.1) and Bacteriophage sp. isolate 1201_113983 (Accession Number: OP030905.1). Average nucleotide identity (ANI) values between isolated Klebsiella phages MFS and other phages ranged from 81% to 97% (as shown in Table 1), with Klebsiella phage PWKp20 being the closest previously characterized bacterio-phage according to ANI values, isolated in Sweden. To obtain a maximum likelihood phylogeny, a multiple whole-genome sequence alignment of Drulisvirus and *Klebsiella* phages πVLC1-4 (58,184 positions) was utilized. This phylogeny indicated that phages πVLC1-4 formed a mono-phyletic group (as depicted in Fig 2).

**Fig. 2.**
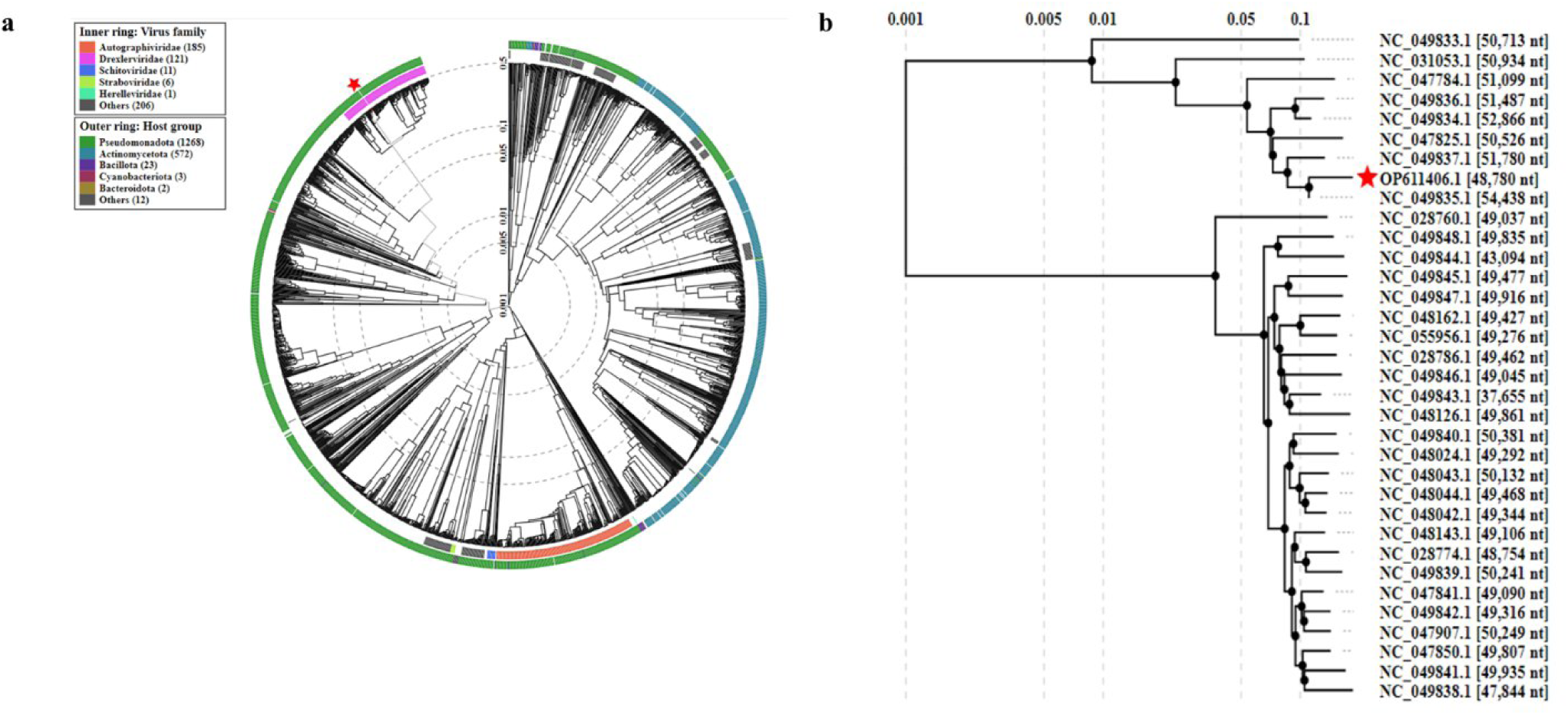
Phylogenetic analysis of phage MFS genome. (a) Circular phylogenetic tree based on proteomic analysis generated by VIPTree. Red stars represent *Klebsiella* phage MFS detected in this study. Besides our *Klebsiella* phage MFS, this tree includes 1922 dsDNA viruses deposited on the VIPTree database, classified by their family (inner ring) and their respective host taxa (outer ring) (b) Rectangular phylogenetic tree (subset) showing the distinct clusters of 34 Klebsiella phages including phage MFS generated using ViPTree, with the log scale on top representing the SG values.

### Bioinformatics Analysis

Nucleotide sequences were compared to those in GenBank using BLASTn and BLASTp (Supplementary File 2). A BLASTn of the assembled long read found the genome to be 96.78% similar to Klebsiella phage PWKp20 (Accession Number: MZ634351.1). The genome had an average of 96.34% ANI with other *K. pneumoniae* phages. High pairwise nucleotide identities (from 93% to 97%) were also observed. This shows the genome as a close relative of the *K. pneumoniae* phages. According to the Phylogenetic analysis of phage proteomes using ViPTree, the phage MFS forms a cluster with other *Klebsiella* phages of Drexleviridae family (Fig 2) and most closely related to *Klebsiella* phage vB_KpnS_Domnhall (NC_049835.1).

The pairwise genome comparison shows that the phage MFS and its closet neighbor *Klebsiella* phage vB_KpnS_Domnhall (NC_049835) have similar genetic make-up, although the location and/or orientation of the genes differs (Fig 3).

**Fig. 3.**
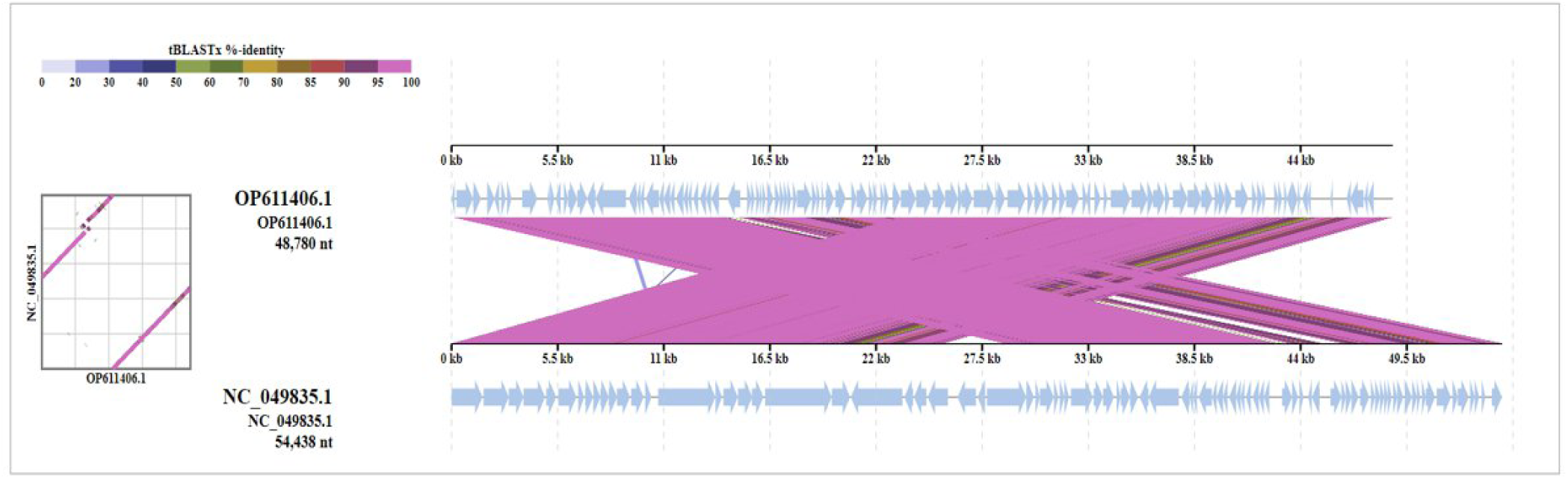
Comparative genomic analyses of *Klebsiella* phage MFS and its closest relative *Klebsiella* phage vB_KpnS_Domnhall (NC_049835). The different colors of shading below each genome indicates sequence similarities between the genomes.

### Genome Annotation

A total of 119 coding DNA sequences (CDSs) encoding polypeptides were found in the MFS genome. All predicted protein-coding genes were screened using BLASTP and Psi-BLAST algorithms against the no redundant protein database at NCBI. From the 119 coding sequences (CDSs), 43 have assigned functions, and 76 were predicted to be hypothetical proteins. The functional genes are highlighted on the genomic map (Fig 4). Of the 119 CDS, 90 were transcribed on the positive strand, and 29 were on the reverse strand. Further information, such as the positions, directions, and putative function of each CDSs, are mentioned in the Supplementary File 1 S2. A total of 41 rho–independent transcription terminators sites were also predicted (Supplementary File 1 S4) in the phage MFS genome. The CDS encode structural proteins (major capsid proteins, tail fiber proteins), host lysis (holin, endolysin and spanin) and proteins involved in other functions (replication, transcription and its regulation, repair and packaging).

**Fig. 4.**
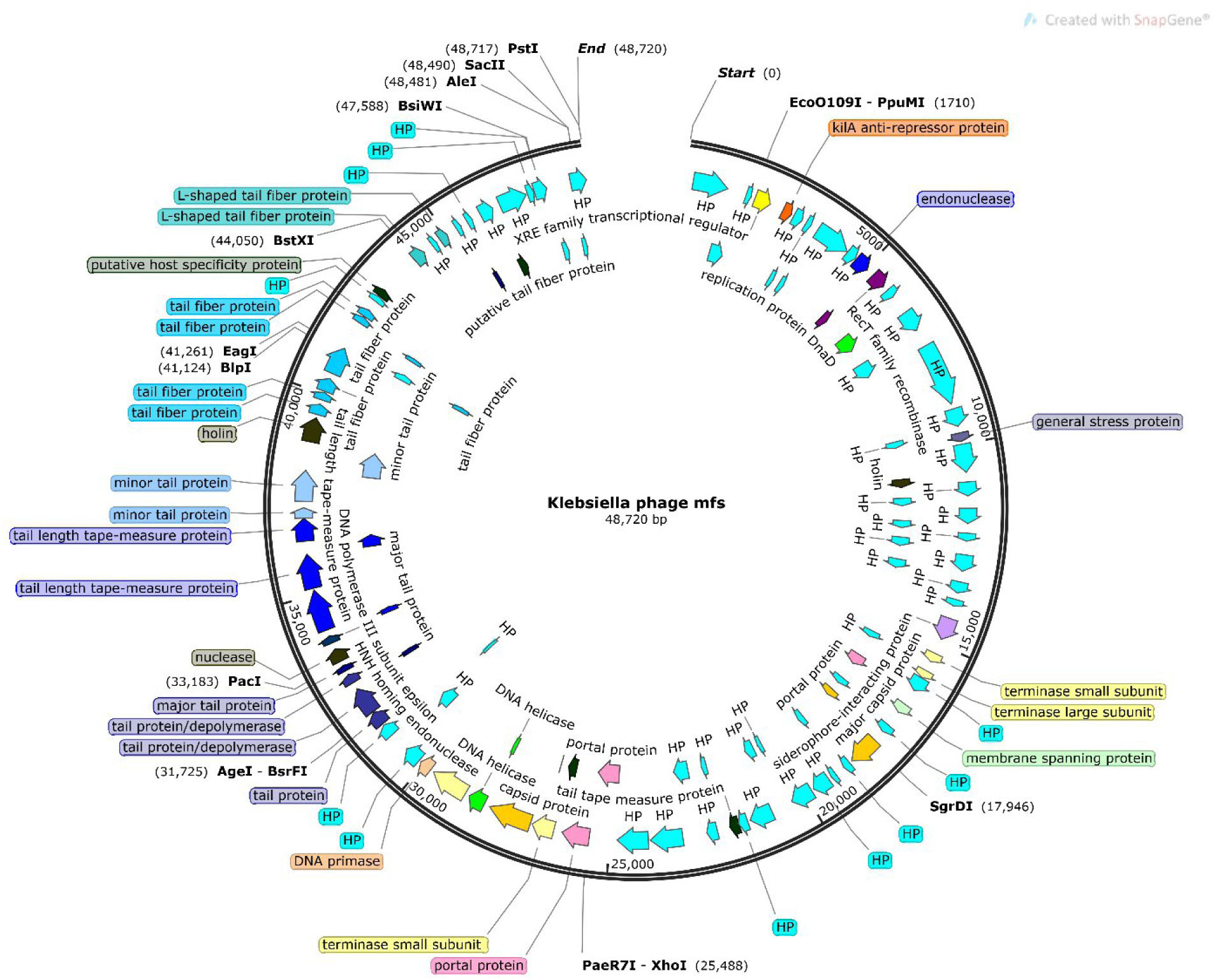
Gene map of *Klebsiella phage* MFS. Different coloured arrows depict predicted CDSs coding different functions: green, DNA packaging; cyan, hypothetical protein (hp); purple/pink, DNA replication, transcription, and metabolism function; black, phage structure; blue, tail structure. The genome map was constructed using SnapGene software.

Our analysis focused on DNA metabolism and replication proteins in phage genomes, identifying key enzymes and their functions. These include a putative polynucleotide kinase/phosphatase (CDS 13), DNA adenine-methyltransferase (CDS 5), DNA helicase (CDS 2), phage-associated recombinase (CDS 117), and a putative single-strand DNA binding protein (CDS 115). These proteins play crucial roles in DNA repair, replication, and recombination, facilitating the phage’s adaptation and survival within host cells.

The 3’-phosphatase, 5’-polynucleotide kinase is a member of the same family as the C-terminal domain found in the bifunctional enzyme T4 polynucleotide kinase/phosphatase PNKP. Within this family, the phosphatase domain of PNKP has the ability to catalyze the hydrolytic removal of the 3’-phosphoryl group from DNA, RNA, and deoxynucleoside 3’-monophosphates [34].

N-6-DNA adenine-methyltransferase, involved in DNA methylation for exonuclease protection, shows high sequence identity with enzymes from Klebsiella phages, suggesting a common strategy among phages to evade host restriction-modification systems [35] The presence of these enzymes indicates the mechanism to escape restriction–modification by a bacterium and also a feature associated with polyvalent phages [36]. The putative helicase and phage-associated recombinase are essential for DNA replication and horizontal gene transfer, respectively, underpinning the phage’s evolutionary capabilities [37].

Our analysis also revealed the presence of putative exonucleases with high sequence identity to those in Klebsiella phages, involved in DNA degradation and stability maintenance. Moreover, a DNA-binding transcriptional regulatory protein (CDS 42) and a single-strand DNA binding protein (CDS 115) were identified, highlighting their roles in transcription regulation and DNA replication.

In summary, the identified enzymes—polynucleotide kinase/phosphatase, recombinase, exonuclease, and transcriptional regulatory proteins—are integral to the DNA metabolism and recombination processes in phages, facilitating efficient replication and adaptation within host cells. This insight into phage biology underscores the complex interactions between phages and their host bacteria, offering potential avenues for therapeutic applications.

### Functional Analysis

In the phage MFS genome, several coding sequences (CDS) are instrumental in morphogenesis and DNA packaging. For instance, CDS.68 encodes a major capsid protein with full identity to its counterpart in Klebsiella phage PWKp19 and nearly full identity to that in *Klebsiella* phage vB_KpnS-VAC6. Proteins likely involved in tail formation are encoded by CDSs 79, 80, 81, 89, and 90, while CDSs 91 and 92 are implicated in tail assembly. Specifically, CDS 81’s product, a major tail protein, has significant identity to depolymerases in other *Klebsiella* phages, highlighting its role in breaking down host capsules.

Further analysis identified genes (CDSs 93 to 99) hypothesized to encode tail fiber proteins, crucial for phage specificity and host range due to potential mutations altering their structure. Tail length determination proteins, or tape measure proteins, are encoded by CDSs 84 to 88, with a combined sequence forming a protein that closely matches those in Klebsiella phage KOX1 and PWKp19, suggesting a tail length of approximately 145.35 nm based on the correlation between amino acid count and physical length. The tape measure protein dictates the tail length and enables DNA transit to the cell cytoplasm during infection [38].

Key DNA packaging elements include the small and large terminase subunits, encoded by CDSs 61, 62, and 63, critical for ATP-dependent DNA packaging into the phage capsid. This system underlines the complex interplay of structural proteins in phage assembly and host infection mechanisms [39]. By distilling essential genes and their functions within the MFS phage genome, we underscore the sophisticated architecture governing phage infectivity and replicaion, providing insights into their potential manipulation for therapeutic and biotechnological applications.

The absence of coding sequences for excisionase, integrase, or repressor genes in the genome of phage MFS indicates that the phage MFS has a lytic life cycle. The culmination of the phage lytic cycle involves the breakdown of the bacterial cell wall, leading to the release of progeny phages. This process typically involves two phage-encoded proteins: a pore-forming protein known as holin and a cell wall-degrading enzyme, either phage lysozyme or endolysin. The holin-endolysin system is present in Almost all dsDNA phages. Holin, a small protein with a transmembrane domain, makes a pore through which endolysin reaches the bacterial cell wall and cleaves ß1,4-glycosidic linkage of the peptidoglycan layer of bacterial cell walls. The canonical order of lysis genes in a phage of Gram-negative hosts is holin, endolysin and spanin. We observe the same orientation of three coding sequences of the lysis cassette. The phage MFS genome contains three CDS 16, CDS 17, CDS 18 that encode putative holin, putative endolysin and putative spanin, respectively. The presence of lytic components verifies the results of clear plaque formation in the DLA method [40, 41].

A putative holin encoding gene CDS 16 is located upstream of CDS 17, which encodes a putative endolysin. The small size (71 amino acids) of the protein encoded by CDS 16, prediction of one transmembrane α-helical domain and a large periplasmic domain indicates that the MFS holin belongs to class III holin similar to that and found in T4-like and T5-like phages [42]. Moreover, DeepTMHMM predicted the presence of a signal peptide from the N-terminal end (aa 1-19) (Fig 5).

**Fig. 5.**
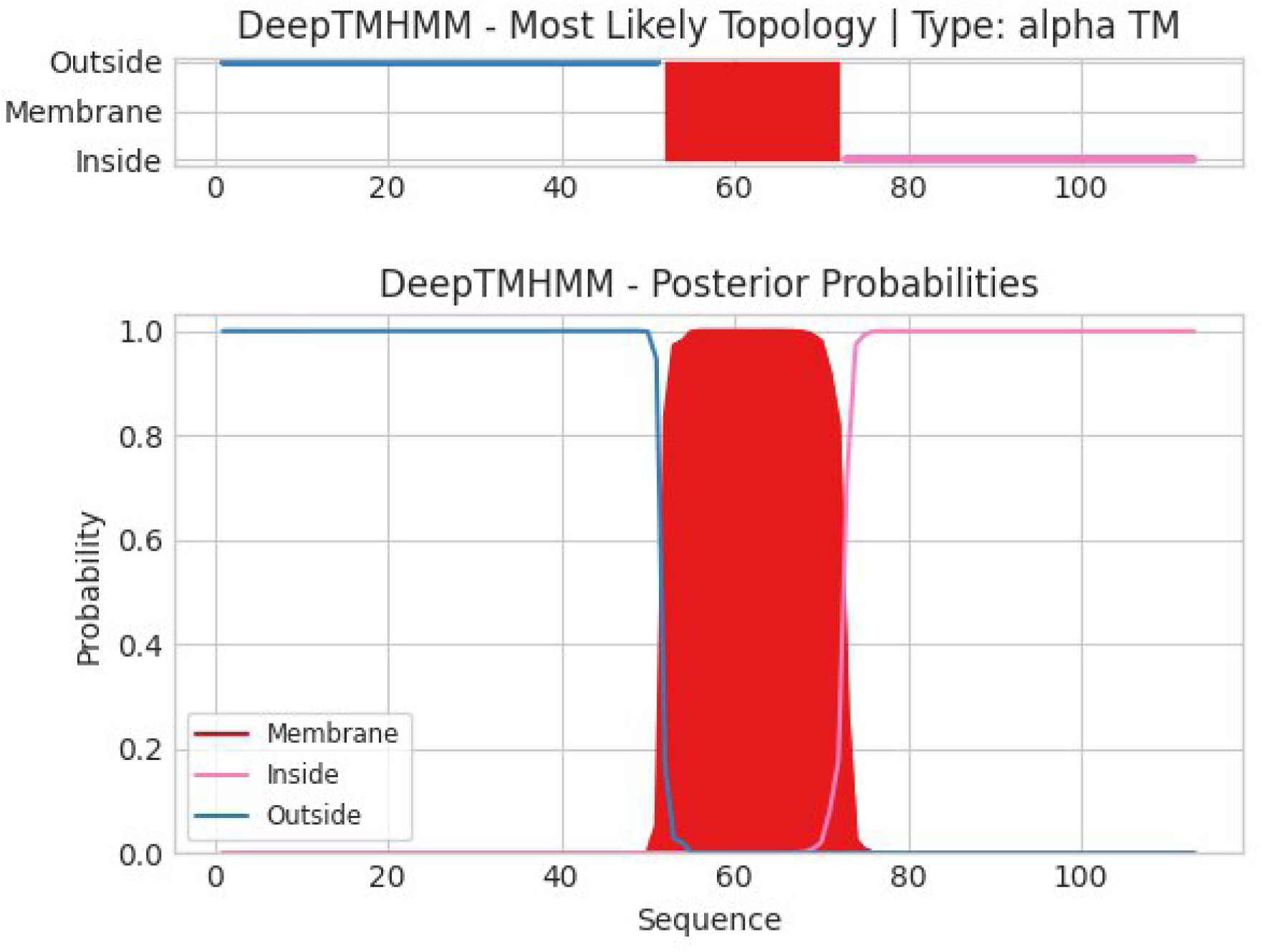
DeepTMHMM – Predictions of CDS 16 (putative holin)

The putative endolysin (CDS 17) of phage MFS is predicted to be a SAR-endolysin. Motif analysis showed that this putative endolysin has similarity with R21 and LyzP1, the lysozymes of coliphages 21 and P1 and, respectively. R21 and LyzP1 were the first endolysins shown to have N-terminal SAR (for signal-anchor-release) domains [43].

Analysis of CDS17 reveals few important features: at the N-terminal region a short stretch of 4 to 6 amino acids with a net positive charge (lysine and arginine residues), which function as a positive anchor to the negatively charged inner side of the cytoplasmic membrane, a H-region(hydrophobic region) is composed of hydrophobic residues and tends to form an a-helix, enhancing insertion of the signal peptide into to phospholipid double layer, CDS 17 has N-terminal region rich in hydrophobic amino acids albeit compared to other SAR-endolysins, this region does not contain glycine or serine residues, but contains alanine, valine and isoleucine. In addition, immediately after the H-region the conserved catalytic glutamate residue (E24) is present. The characteristic catalytic triad motif is composed of Glu 24, Asp 33 and Thr 39. Amino acid residues conserved in other endolysins are also present, the salt bridge forming residues Glu 24 and Arg 147 are also present in the amino acid sequence (Fig 6).

**Fig. 6.**
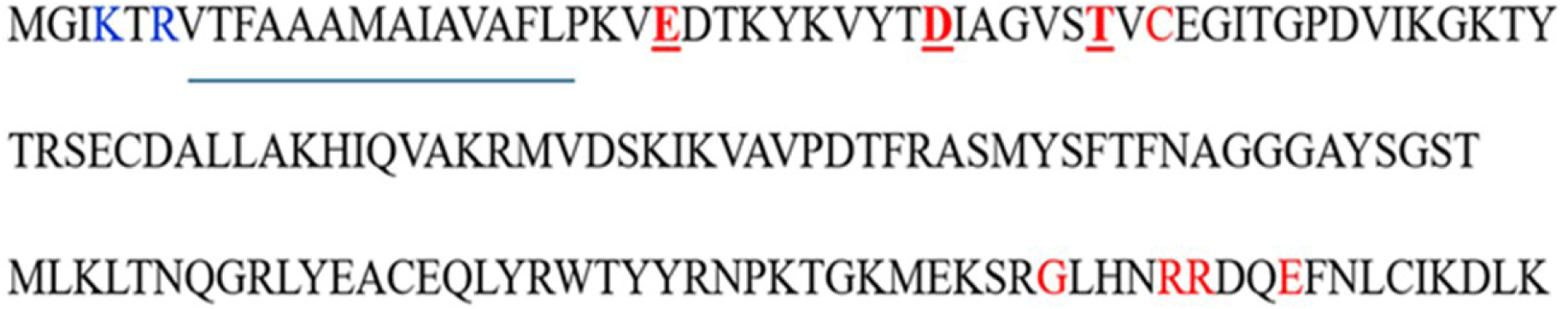
The amino acid sequence of CDS17, putative SAR-endolysin of phage MFS. The basic residues at the N-terminal region are highlighted in blue, the hydrophobic region is underlined, the conserved residues are highlighted in red and the catalytic triad are highlighted in red, bold and underlined.

The endolysins of bacteriophage P1 (Lyz) and coliphage 21 (R21), are orthologs of T4 lysozyme [44, 45], But due to presence of an N-terminal transmembrane domain (TMD) that act as a type II signal anchor, or uncleaved signal peptide, these enzymes are exported by the bacterial sec system and anchored to the membrane by the N-terminal SAR domain [46]. This is in contrast to the canonical phage lysozymes, such as the one in phageT4, which accumulate in a fully folded and enzymatically active state in the cytoplasm. The loss of the proton motive force (PMF) due to the pinholin, causes the release of the tethered SAR endolysin, which then folds into an active state to degrade the peptidoglycan layer in the cell wall [43].

The CDS 18 of phage MFS encodes a putative spanin. BLASTp search showed that it has 99% identity with Rz-like spanin [*Klebsiella* phage *KOX1*] and [*Klebsiella* phage vB_KpnS_KpV522. Further analysis revealed that the putative spanin has a lipopeptide signal peptide at the N-terminal end, a non-cytoplasmic or periplasmic region followed by a transmembrane domain and a cytoplasmic domain (Fig 7). The features match with those of U-spanin of *Escherichia coli* phage T1 (T1 phage). U-spanins are grouped into 13 families. T1gp11 is the representative member of the largest family. In the periplasmic region, the predicted secondary structures were conserved across all members, that is a short stretch of alpha helices followed by 4 or more beta sheet elements. The predicted secondary structure of CDS-18 showed the same pattern of structural arrangement (Fig 6). Except for J8–65 and Limezero, all other members of the T1gp11 u-spanin family were part of pinholin and SAR-endolysin lysis cassettes [46]. We have predicted the lytic machinery of the Phage MFS also consists of pinholin and SAR-endolysin. Therefore, we predict that CDS-18 of phage MFS is a U-spanin. U-spanins are thought to disrupt the outer membrane by fusing the inner and outer membranes of the host cell. This process likely involves conformational changes mediated by beta-sheets, similar to how type II viral fusion protein’s function [47, 48].

**Fig. 7.**
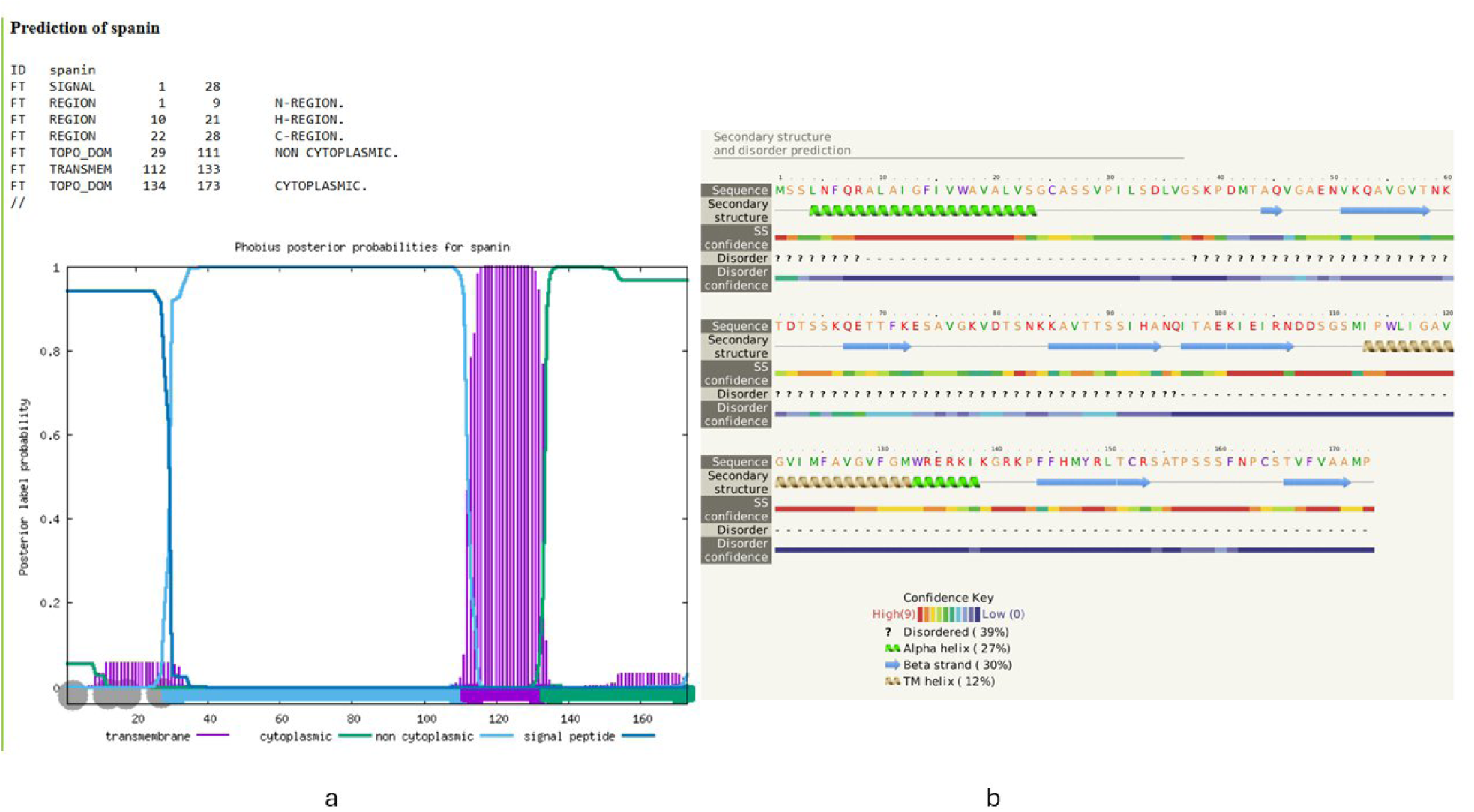
The prediction of the Transmembrane domain and secondary structural elements in CDS 18 (putative U-span of phage MFS) was carried out by Phobius (a) and by Phyre2 (b), respectively

Through computational analysis, including GC-skew plotting, the probable origin of replication in the MFS phage was identified near nucleotide 13100, adjacent to CDS 32, providing insights into the phage’s replication mechanisms.

Additionally, 76 genes of unknown function were identified. The hypothetical proteins investigated through BLAST search analysis provided information about the putative functions of 19 CDSs, which reflect the diverse functions, including roles in structure, enzymatic activity, transcriptional regulation, and membrane function (Supplementary File 1 S3). Sequence homology was found between some hypothetical proteins of MFS and phage proteins encoded by different bacteriophages of *K. pneumoniae*.

Furthermore, no toxin-, virulence factor-, or antibiotic resistance-related genes were found, indicating that MFS is a virulent and potential candidate for phage therapy. The genome of phage MFS contains numerous hypothesis proteins of unknown function; therefore, further comprehensive functional analysis is required to determine the safety of using these phages in therapeutic applications.

## Discussion

*K. pneumoniae* has emerged as a significant antibiotic-resistant (AMR) pathogen in healthcare and community settings. As antibiotic resistance poses a significant threat to public health, the use of bacteriophages to treat pathogenic bacterial infections has recently gained attention. Research has explored the use of lytic phage therapy to target specific multi-drug resistant (MDR) bacterial ESKAPE pathogens, such as *Pseudomonas aeruginosa*, *Enterococcus spp. Staphylococcus aureus*, *Acinetobacter baumannii*, and *Escherichia coli* [49,50,51,52,53,54,55,56]. Therefore, isolating and conducting comprehensive studies on phages and their host communities is crucial before considering bacteriophages as a viable solution for combating bacterial pathogens.

The main aim of this study was the isolation and characterization of a lytic bacteriophage with potential for prophylactic/therapeutic use against MDR and XDR *K. pneumoniae*. We investigated the genomic properties of a newly isolated bacteriophage MFS which infects clinical *K. pneumoniae* strains. The lytic activity exhibited by phage MFS against various MDR and XDR *K. pneumoniae* strains underscores its potential as a therapeutic agent. Producing clear plaques by *Klebsiella* phage MFS indicates robust lytic capabilities, essential for effective phage therapy [57]. Additionally, genes encoding endolysin and holin reinforce its lytic nature, suggesting that phage MFS can efficiently disrupt bacterial cell walls and lyse the host bacteria [58].

Genomic analysis of phage MFS revealed a genome size of 48,780 bp, comprising 94 protein-coding sequences, which include essential genes for phage replication, structure, and host lysis [59]. Notably, the genome lacks genes associated with antimicrobial resistance, virulence, and lysogeny, indicating its suitability for therapeutic applications [60]. This absence reduces the risk of transferring undesirable traits to bacterial hosts, thereby enhancing the safety profile of phage MFS for clinical use [61].

This study highlights the potential of hospital sewage as a valuable source of bacteriophages. The isolation of phage MFS from such an environment suggests that similar sources could be exploited to discover phages targeting other clinically significant pathogens [62]. The high sequence similarity between phage MFS and other *Klebsiella* phages, as shown by BLAST and phylogenetic analyses, suggests a conserved mechanism of action, which could be harnessed to develop phage cocktails aimed at a broader range of bacterial strains [63].

Despite the promising outcomes, several challenges need to be addressed before phage therapy can be widely adopted. Although beneficial for targeted therapy, the specificity of phages requires comprehensive characterization of phage-host interactions and the natural diversity of phages [64]. Furthermore, the potential for bacterial resistance to phages requires continuous monitoring and possibly the formulation of phage cocktails to minimize resistance development [65].

Further research should prioritize in-depth genomic and physiological characterization of the phage. Employing advanced techniques such as Pulse Field Gel Electrophoresis (PFGE) and Transmission Electron Microscopy (TEM) can offer valuable insights into the genetic composition and structural attributes of the phage [66]. Moreover, assessing the therapeutic effectiveness of phage MFS in animal models would represent a crucial advancement towards clinical implementation [67].

## Conclusion

In conclusion, the isolation and characterization of *Klebsiella* phage MFS from hospital sewage underscore the potential of environmental samples as reservoirs for therapeutic bacteriophages. The phage’s strong lytic activity and the absence of harmful genes position it as a promising candidate for phage therapy against MDR/XDR *K. pneumoniae* infections [57, 60]. Continued exploration and characterization of phages from diverse environments could lead to innovative treatments addressing the growing threat of antibiotic-resistant bacterial infections [61, 62].

## Data Availability

All data produced in the present work are contained in the manuscript

## Ethical statement

The study was conducted following guidelines and approved by the North South University Institutional Review Board (2020/OR-NSU/IRB/1205).

## Availability of data and materials

All of the data are included in the article.

## Data Summary

The genomic sequence of the novel phage has been deposited in GenBank under accession number: **OP611406**. All data from the analyses carried out in this study are available in the Supplementary data files.

